# Nutritional status and its associated factors among HIV positive adolescents on Atazanavir-based regimen attending an urban clinic in Uganda

**DOI:** 10.1101/2020.10.04.20206722

**Authors:** Dave Darshit, Ainembabazi Provia, Nana Nakiddu, Erin Sodawasser, Katrina Harper, John M. Ssenkusu, Sabrina Bakeera-Kitaka, Melanie Nicol, Joseph Musaazi, Christine Sekaggya-Wiltshire

## Abstract

**Background:** Adolescents between the ages of 10-19 represent a growing portion of people living with HIV worldwide. A large proportion of adolescents living with HIV suffer from severe malnutrition because of the chronic ill health and this has been associated with increased morbidity and mortality particularly in Sub-Saharan Africa. Little is known about the nutrition status of adolescents living with HIV who are on second line treatment particularly Atazanavir. Therefore, we assessed the nutrition status and associated factors among HIV positive adolescents on Atazanavir-based regimen attending an urban clinic in Uganda.

**Method:** This was a cross-sectional study carried out between December 2017 and July 2018. Using convenience sampling, adolescents aged 10-19 years attending an urban clinic in Kampala on Atazanavir-based regimen were enrolled into the study. Nutritional status was assessed using BMI-for-Age and Height-for-age as measures of thinness and stunting respectively. Standard deviation scores (Z scores) were applied to determine the nutritional status. WHO and CDC Z-score cut offs were used to categorize the nutrition status. Data was entered into an electronic database using REDCap. Statistical analysis was done using STATA version 15.1 (Texas, USA).

**Results:** Data from 132 adolescents were included. We found that 28% were malnourished (composite outcome of stunting and thinness). The prevalence malnutrition of thinness was 7.6% with 2.3% being severely thin. The prevalence of stunting was 23.7% with 1.5% being severely stunted. Adolescents with no parent were more likely to be malnourished than adolescents who had either one or both parents (Adjusted Odds Ratio [AOR]: 3.70 95% Confidence Interval [CI]: 1.20-11.37, p=0.023). In addition, adolescents who had attained at least secondary education were less likely to be malnourished (AOR: 0.40, CI:0.17-0.95, P-value=0.037).

**Conclusion:** There is a high proportion of adolescents with HIV who are malnourished. Low level of education (No education and elementary) and having no parent are important risk factors to malnutrition in this population. There is need for optimizing nutrition care for adolescents on HIV treatment.

## Introduction

Adolescents between the ages of 10-19 years represent a growing portion of people living with HIV worldwide (1). Globally, 1.7 million adolescents were living with HIV in 2019 accounting for 5% of all the people living with HIV(1). Majority (88%) of these adolescents are from sub Saharan Africa(1). In Uganda approximately, 1.5 million people were infected with HIV as of 2019 with a 5.8% prevalence amongst people aged between 15-49 years(2). Majority (85%) of whom were on treatment and more than half having their viral load suppressed(2). According to World Health Organization, about 94.1% and 5.4% of patients were estimated to be receiving first line and second-line treatment respectively (3). It was also estimated that the proportion of people receiving first line therapy would decrease to 94.1% as those receiving second line therapy increase to 5.9% (3) due to increase in prevalence of drug resistance to first line regimen estimated between 3-29% (4). This denotes the importance of addressing major threats to people living with HIV, which may affect HIV treatment response and outcomes as they advance through treatment lines. Malnutrition is a common problem amongst people living with HIV and has been linked to treatment failure(5), increased morbidity and mortality (6, 7). The physical and psycho-social changes occurring during childhood and adolescence (8) makes adolescents more vulnerable to health and nutrition concerns compared to others. For example, the total nutrient needs of adolescents are higher than at any other time in the human lifetime (9). The Sub-Saharan African region has had a high burden of malnutrition amongst children and adolescents (10-12). In Uganda, approximately 36.2% of HIV positive adolescents receiving care are stunted with 11.1% being severely affected (13).

The role played by nutrition in influencing the somatic, intellectual and social aspects of HIV-exposed children has been reported (11). Additionally, the HIV infection and drugs may compromise nutrients intake as well as increasing nutritional demand required through several metabolic processes (14). Of particular importance is the Atazanavir (ATV) -based combination that is one of the preferred proteases inhibitors (15, 16) for clinical use in our setting. Atazanavir has shown to induce nausea in 4-14% of the population and diarrhea 3-11% of the population similar to other protease inhibitors (17). These adverse drug events may influence dietary uptake as well as nutrient bioavailability in the body. Also, malnutrition may affect the bioavailability of drugs in the body (18)

Despite the fact that adolescents are prone to malnutrition, inadequate research has been conducted to inform adolescent health agenda particularly in the context of second line HIV treatment and care. We assessed the nutritional status of adolescents receiving ATV-based regimen attending an HIV outpatient clinic in an urban setting in Uganda.

## Materials and Methods

### Study design, site and population

This was a cross-sectional design study conducted between December 2017 to July 2018 among HIV positive adolescents on ATV based regimen aged between 10 to 19 years attending the Pediatrics Infectious Diseases Clinic. The Pediatrics Infectious Diseases Clinic is managed by Baylor College of Medicine, Kampala, a leading provider of family centered child and adolescent HIV/AIDS services within Kampala, Uganda. At the time of the study, there were 3326 HIV positive adolescents enrolled in care with 597 on second line therapy, of which 317 adolescents were on ATV based Anti-Retroviral Treatment (ART).

Adolescents who had been on ATV based antiretroviral regimen for at least 6 weeks prior to enrolment and consented or assented to participate with the parents’ consent were enrolled into the study. The adolescents who had known history of liver failure and end stage renal disease or cardiovascular disease which on their own could lead to malnutrition were excluded out of the study.

### Study procedures

We enrolled adolescents using cconvenience sampling procedure. Convenience sampling was used because our study population was small, and the adolescent specific clinic visits were twice a week and thus enrolled any patient aged 10-19 years on ATV based ART regimen that we came across. Adolescents were referred to the study team after the clinic visit procedures by the attending Health care worker. The adolescents aged 18 and 19 consented while for adolescents aged 10-17 years were assented and their parents consented before participating in the study. The adolescents were assessed for understanding of the study procedures and later interviewed. The interview took about 25 Minutes. Adolescents who did not have time to participate in the study after the clinic procedures were called back on appointment for study procedures on the next convenient day.

### Data collection

A structured pretested interviewer-administered questionnaire in English and Luganda languages were used to collect data. The questionnaire consisted of socio-demographic information and relevant clinical history of interest including duration on ART and the ART regimen. Adherence was measured using a self-report tool (19). The self-report tool had stemmed questions, evaluating challenges in adhering to ART: (a) Do you sometimes find it difficult to remember to take your medicine? (b) When you feel better, do you sometimes stop taking your medicine? (c) Thinking back over the past four days, have you missed any of your doses? (d) Sometimes if you feel worse when you take the medicine, do you stop talking it?

Weight was measured using an electronic Seca scale whose calibration was done on a daily basis. Body weight was measured with minimal clothing (Excluding shoes, belts, socks, watches and jackets) and recorded to the nearest 0.1 kg.

Height measurements were taken at maximum inspiration using a standardized erect stadiometer recorded to the nearest 0.1 cm.

Pharmacokinetics samples were taken from the 16 malnourished and 19 randomly selected well-nourished participants at 22-24 hours after the last dose and ATV trough concentrations (mg/ml) were analyzed using a High-Performance Liquid Chromatography at the Infectious Diseases Institute translation laboratory.

Viral load performed nearest six months before enrollment into the study was extracted from the electronic medical database.

### Data management and analysis

We used REDCap ® to collect the data. The data from REDCap was exported to STATA version 15.1 (Texas, USA) for analysis. The nutrition status of adolescents was assessed using BMI-for-Age (BAZ) and Height-for-age (HAZ) indices. Standard deviation (SD) scores (Z scores) were applied to determine the nutritional status using the WHO 2007 reference (for children ≥5 to 19 years) computed by WHO Anthro Plus software(20).

Adolescents whose BAZ <-2 were considered to be thin, and those who had a score of ≥ -2 to, ≤ +1 were considered normal weight (well-nourished) while those who scored >+1 were considered overweight. Adolescents whose HAZ was <-2 were considered to be stunted while those who scored ≥ -2 were considered to be of normal stature (well-nourished). Adherence was categorized as high (no to all questions), medium (yes to one question) and low (yes to two or more questions) (19). We used descriptive analysis to describe participant characteristics. Nutritional status indices were presented as frequency and percentages. Predictors of malnutrition were assessed using Chi-square tests at bi-variable and logistic regression at multivariable analysis. Factors were included into multivariable logistic regression if they had P-value<=0.2 at bi-variate analysis. Statistical significance was considered using 95% confidence interval and P-value<0.05.

### Ethical clearance

The study was approved by Baylor College of Medicine Scientific Review committee, University of Minnesota Ethics Committee (STUDY00000414), The AIDs Support Organization (TASO) Research Ethics (TASOREC/33/17-UG-REC-009), and Uganda National Council of Science and Technology (HS100ES). Adolescents 18 years and above provided written informed consent while those aged 8 – 17 years old provided assent while consent was obtained from their parents/guardians.

## Results

### Social demographic and clinical characteristics of participants

Of the 132 HIV positive adolescents enrolled, 65% were males and the mean age (SD) was 17.1(8) years. The majority (89.4%) were late adolescents aged between 15-19 years. The mean duration on ART was 8.8 (SD:3.3). We found 45.8% were not virologically suppressed and 47.7% had a high adherence. The geometric mean for the Atazanavir C_trough_ pharmacokinetic concentrations was 0.71 (95% CI: 0.55-0.92). The mean C_trough_ pharmacokinetic concentrations for the well-nourished adolescents was 0.66 (95%CI: 0.53-1.07) which was no different from the mean C_trough_ concentrations for the malnourished adolescents was 0.66(95%CI: 0.42-1.03). Approximately 45% of the adolescents had virological failure (viral load >1000). Viral load p-value on chi-square was 0.970. Details of demographic and clinical data are summarized in Table 1.

**Table 1:** Social demographic and clinical characteristics of HIV positive adolescents on Atazanavir based ART regimen

| Variable | Categories | Frequency (%) |
| --- | --- | --- |
| Age (in years) | 10-14 | 14 (10.6) |
|  | 15-19 | 118(89.4) |
| Gender | Female | 67(50.8) |
|  | Male | 65(49.2) |
| Biological parents alive | Yes, both | 47(35.6) |
|  | Yes, one | 55(41.7) |
|  | No | 30(22.7) |
| Level of education | None | 4(3.0) |
|  | Elementary | 37(28.0) |
|  | High School | 78(59.0) |
|  | Tertiary | 13(9.9) |
| Schooling status n=112 | Boarding school | 25(22.3) |
|  | Day school | 87(77.7) |
| Nucleotides in ART regimen | TDF | 87(65.9) |
|  | ABC | 31(23.5) |
|  | AZT | 14(10.6) |
| Duration on ART(Years) | 0-5 | 15(11.4) |
|  | 6-9 | 68(51.5) |
|  | 10+ | 49(37.1) |
| Viral Load (copies/ml) n=131 | <1000 | 71(54.2) |
|  | >=1000 | 60(45.8) |
| Adherence (Self-reported) | Low | 28(21.2) |
|  | Moderate | 41(31.1) |
|  | High | 63(47.7) |

### Nutrition status: Malnutrition amongst HIV positive adolescents on ATV based regimen

We found that (37, 28.0%) were malnourished (composite outcome of stunting and thinness) as shown in fig 1. We also found that (7, 5.3 %) were thin, (3, 2.2%) were severely thin and (4, 3.8%) were overweight as shown in fig 2, and (29, 22.1%) adolescents were moderately stunted while (2, 1.5%) were severely stunted as shown in fig 3.

**Figure 1:**
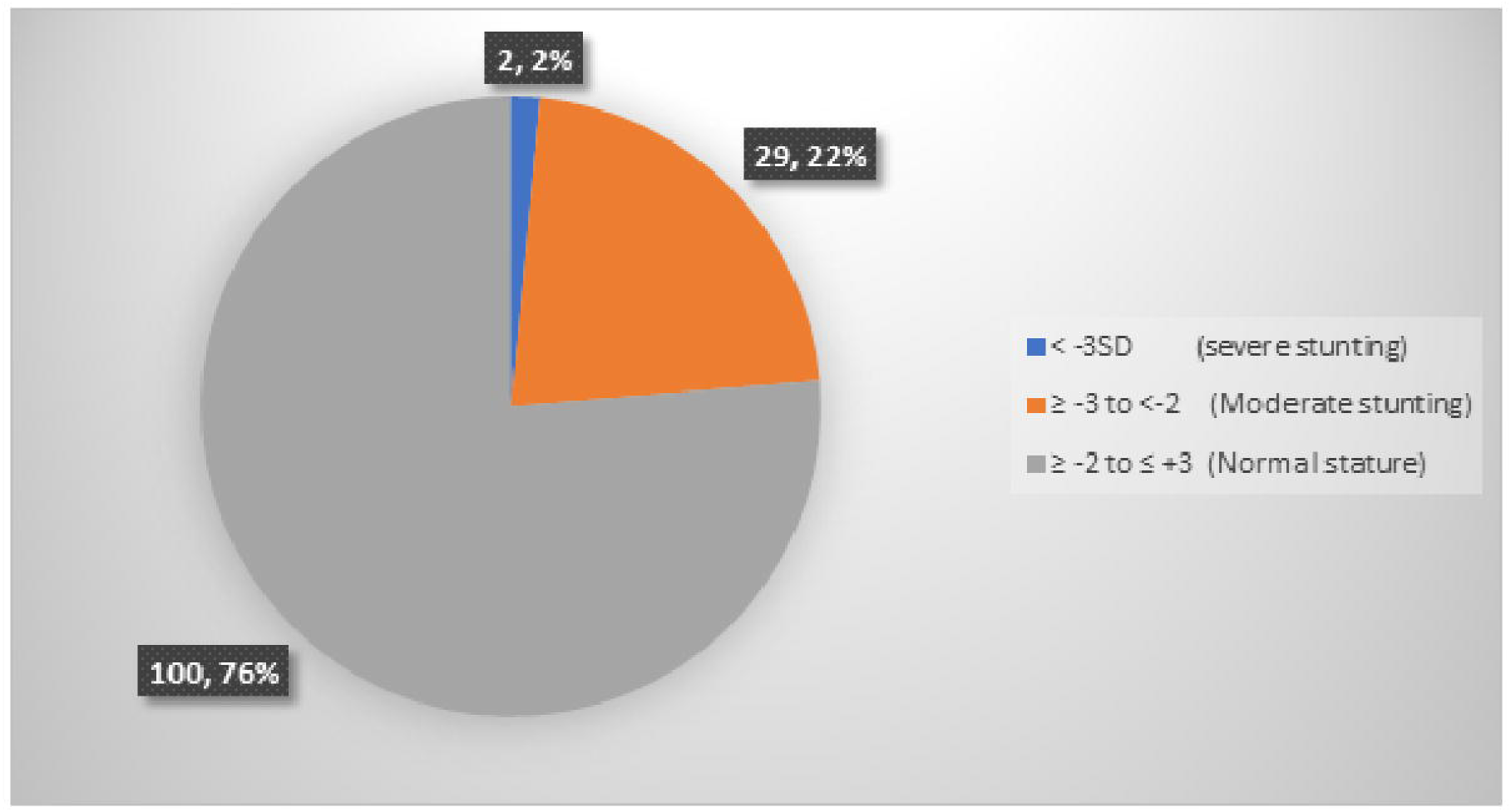
Malnutrition (composite score for thinness and stunting) amongst HIV positive adolescents on ATV based ART regimen

**Figure 2:**
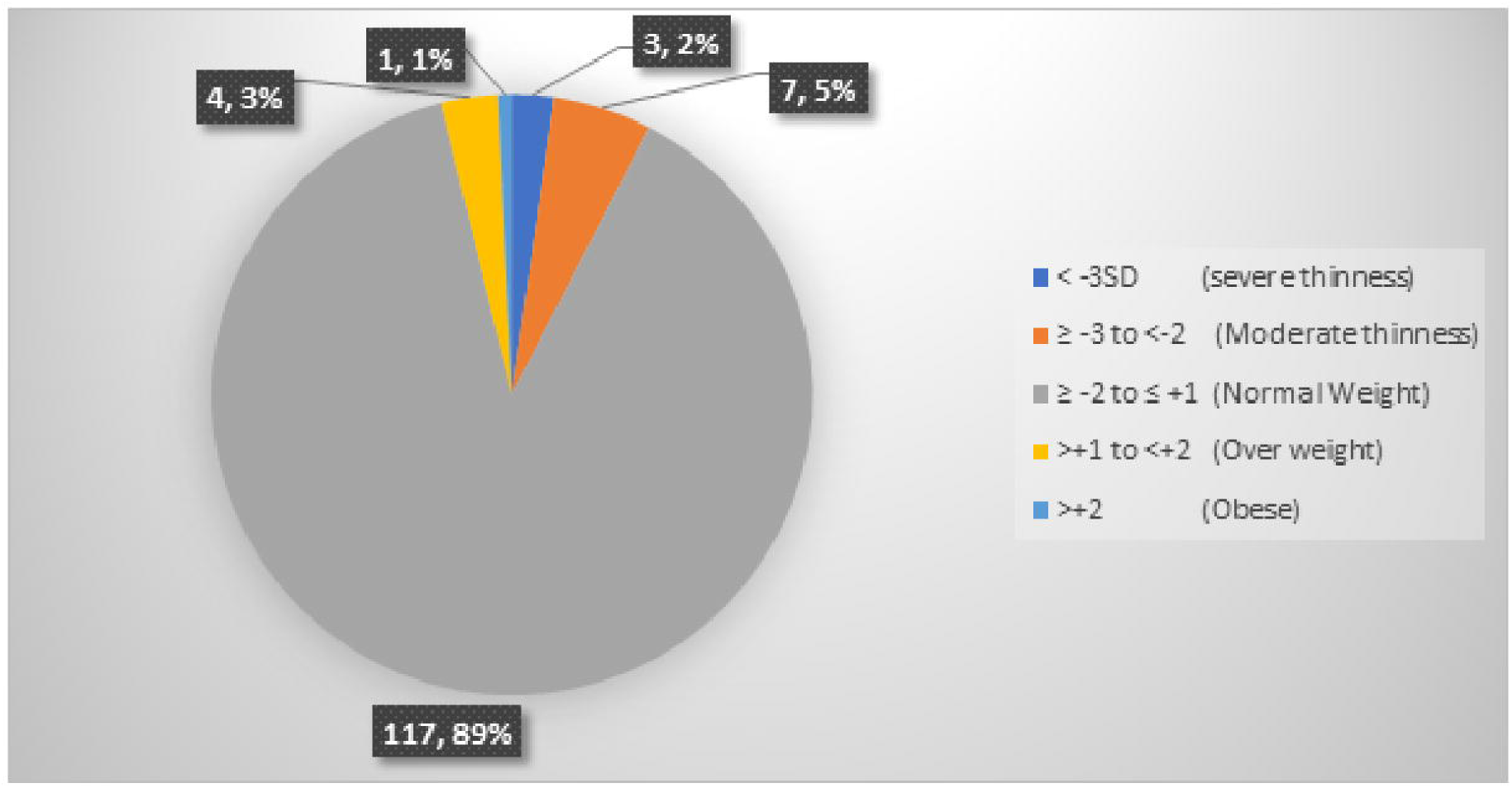
Thinness amongst HIV positive adolescents on ATV based ART regimen

**Figure 3:**
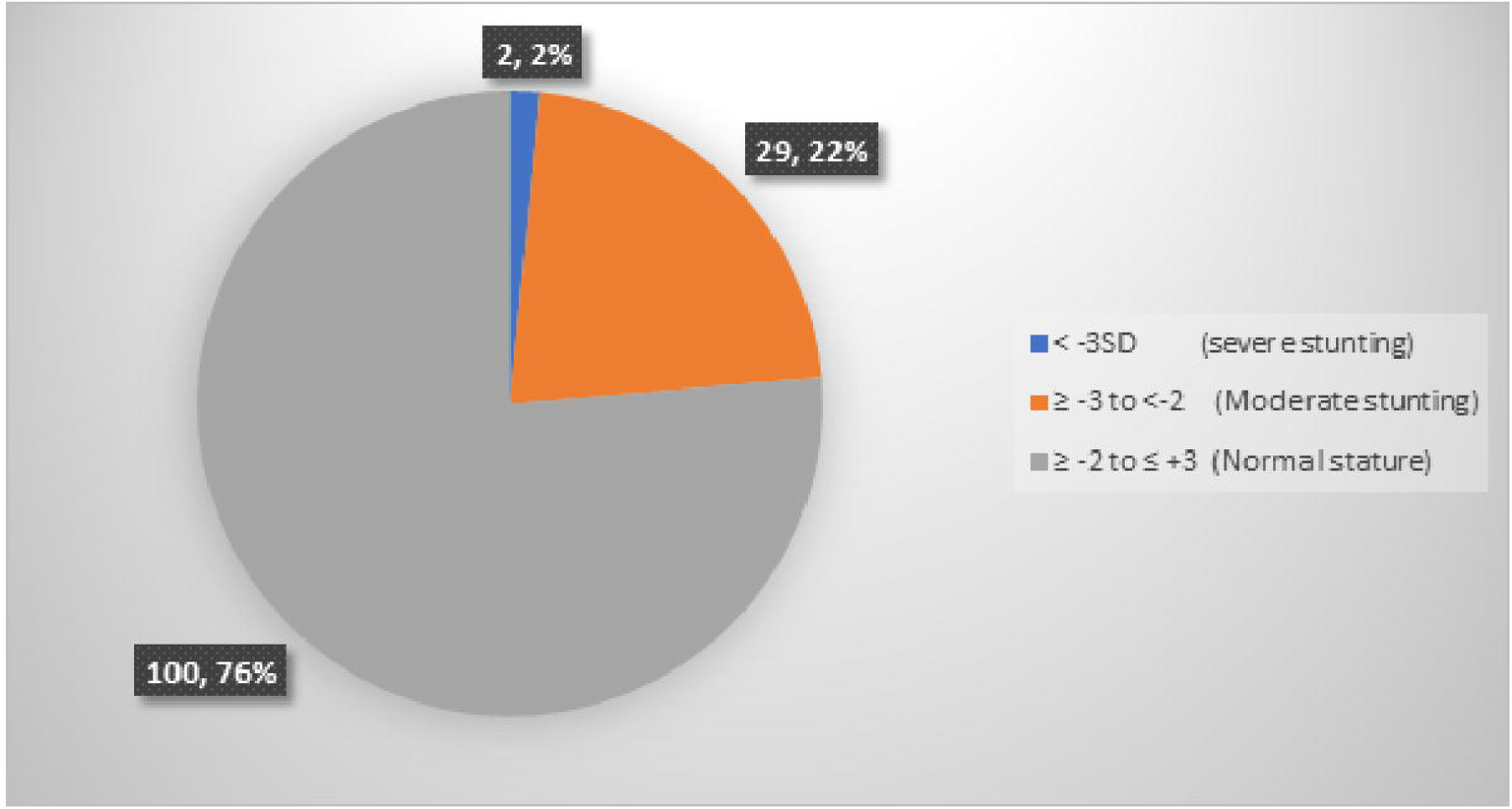
Stunting amongst HIV positive adolescents on ATV based ART regimen

Table 2 shows that adolescents with no parent were more likely to be malnourished than adolescents who had either one or both parents (Adjusted Odds Ratio [AOR]: 3.70, 95% Confidence Interval [CI]: 1.20-11.37, p=0.023). In addition, adolescents who had attained at least secondary education were less likely to be malnourished (AOR: 0.40, CI: 0.17-0.95, P-value=0.037).

**Table 2:**
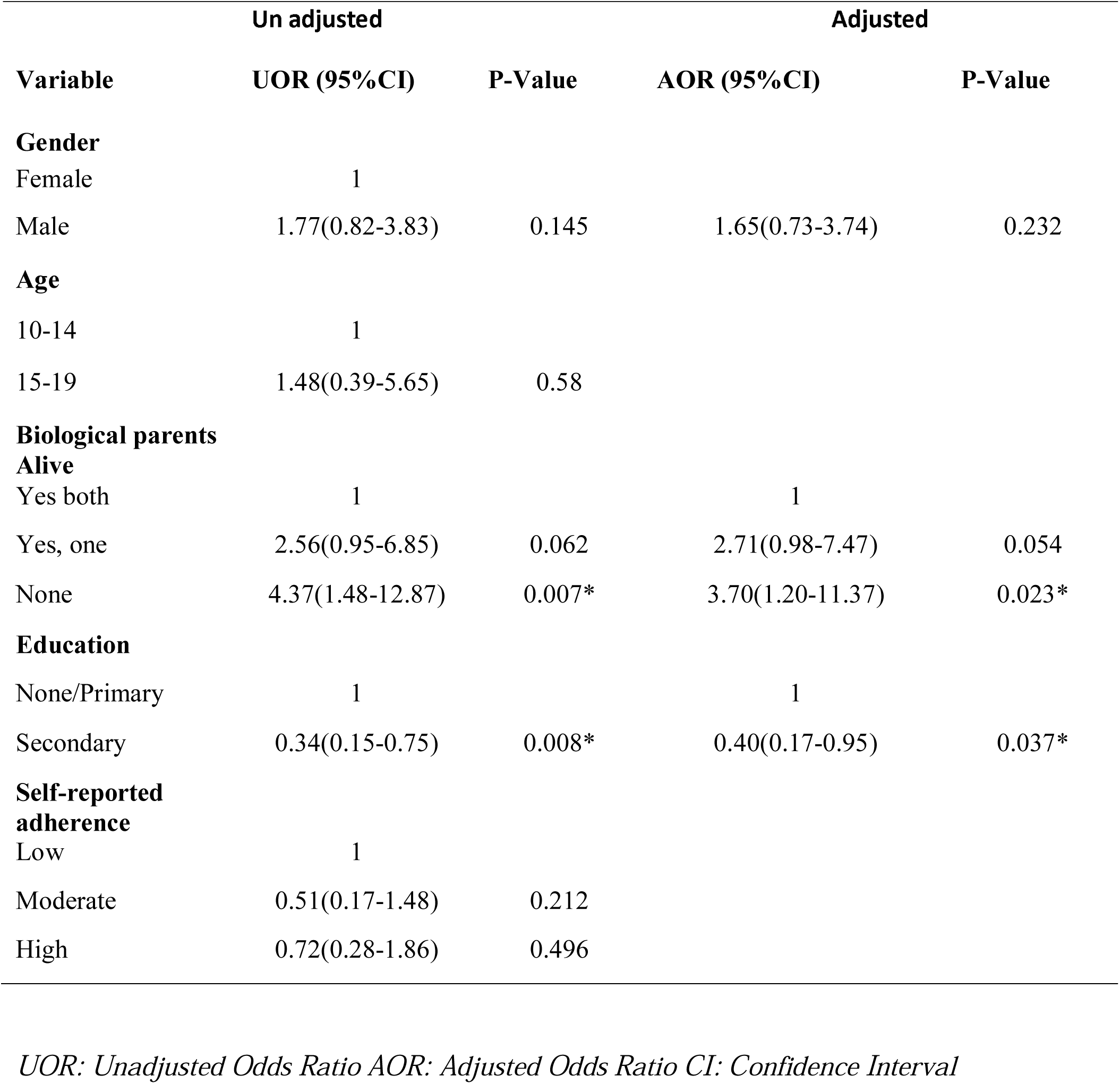
Factors associated with malnutrition (composite outcome of stunting and thinness) amongst HIV positive adolescents on Atazanavir based ART regimen

| Variable | Un adjusted |  | Adjusted |  |
| --- | --- | --- | --- | --- |
|  | UOR (95%CI) | P-Value | AOR (95%CI) | P-Value |
| **Gender** |  |  |  |  |
| Female | 1 |  |  |  |
| Male | 1.77(0.82-3.83) | 0.145 | 1.65(0.73-3.74) | 0.232 |
| **Age** |  |  |  |  |
| 10-14 | 1 |  |  |  |
| 15-19 | 1.48(0.39-5.65) | 0.58 |  |  |
| **Biological parents Alive** |  |  |  |  |
| Yes both | 1 |  | 1 |  |
| Yes, one | 2.56(0.95-6.85) | 0.062 | 2.71(0.98-7.47) | 0.054 |
| None | 4.37(1.48-12.87) | 0.007* | 3.70(1.20-11.37) | 0.023* |
| **Education** |  |  |  |  |
| None/Primary | 1 |  | 1 |  |
| Secondary | 0.34(0.15-0.75) | 0.008* | 0.40(0.17-0.95) | 0.037* |
| **Self-reported adherence** |  |  |  |  |
| Low | 1 |  |  |  |
| Moderate | 0.51(0.17-1.48) | 0.212 |  |  |
| High | 0.72(0.28-1.86) | 0.496 |  |  |

*UOR: Unadjusted Odds Ratio AOR: Adjusted Odds Ratio CI: Confidence Interval*

## Discussion

This study showed that 28% of adolescents receiving second line therapy with Atazanavir for HIV treatment are malnourished (composite outcome of stunting and thinness), and 7.25% of adolescents are thin and 25% are stunted. This prevalence of malnutrition is high and falls short of the strategy to end all forms of malnutrition and address the nutritional needs by 2030 (21).

Our prevalence of malnutrition is lower than what was documented among HIV-infected adolescents aged 10-19 years in rural and urban Uganda 5 years ago where 26% of the adolescents were thin and 45% stunted (13), but comparable to the general population of school-going children aged 6-14 years where 22.5% of the adolescents were reported to be stunted (22). This may reflect the increased access and early initiation of ART in Uganda over time following the test and treat strategy. Higher rates of malnutrition have also been reported in earlier studies conducted in other developing nations amongst HIV infected children and adolescents including Thailand (43 %,), West Africa (44%), India (58%). However, studies in Tanzania and Pakistan have demonstrated slightly lower rates of malnutrition (8-18%) (23).

The variation of malnutrition in different studies might be due to the variable study settings, while our participants attended an urban clinic, some of the comparative studies involved a combination or only rural areas.

Our finding that adolescents with no parents were more likely to be malnourished disagrees with the study from central and West Africa where orphan hood did not have any association with malnutrition (10). Our finding that adolescents who had attained secondary education and above were less likely to be malnourished denotes the importance of education in nutrition care as noted by other studies (24-26).

Malnutrition has not been associated with low Atazanavir concentrations in previous studies which is similar in this population, whereby the C_trough_ concentrations of Atazanavir were comparable in both the malnourished and well-nourished group. However, we were limited by the small sample size and this needs to be evaluated in a larger study.

## Conclusion

There is a high proportion of adolescents with HIV who are malnourished. Low level of education (No education and elementary) and having no parent are important risk factors to malnutrition in this population. There is need for optimizing nutrition care for adolescents on HIV treatment.

## Data Availability

data available on request from the author

## List of abbreviations

ATV: Atazanavir
ART: Anti-Retroviral Therapy
BAZ: Body Mass Index for Age z-scores
HAZ: Height for Age z-scores
HIV: Human Immune-deficiency Virus
WHO: World Health Organization
CDC: Center for Disease Control
SD: Standard Deviation
TASO: The AIDs Support Organization
UOR: Unadjusted Odds Ratio
AOR: Adjusted Odds Ratio
CI: Confidence Interval
TDF: Tenofovir
ABC: Abacavir
AZT: Zidovudine

## Declarations

### Ethics approval and consent to participate

The study was approved by Baylor College of Medicine Scientific Review committee, University of Minnesota Ethics Committee (STUDY00000414), TASO Research Ethics (TASOREC/33/17-UG-REC-009), and Uganda National Council of Science and Technology (HS100ES). Adolescents 18 and 19 years provided written informed consent while those aged 8 – 17 years old provided assent while consent was obtained from their parents/guardians before participating in the study.

### Availability of data and materials

The datasets used and/or analysed during the current study are available from the corresponding author on a reasonable request. All data generated or analysed during this study are included in this published article.

### Funding

The study was supported by the Uganda Research Training Collaboration; a collaboration between Makerere University and University of Minnesota to mentor students in a research career. The funding bodies did not have a role in the design of the study and collection, analysis, and interpretation of data and in writing the manuscript.

#### Authors’ contributions

AP, DD, NJ, ES, MN, SKB and CSW conceptualised the study and wrote the protocol. AP, DD, NJ, ES participated in data collection. MJ and JS were responsible for the data analysis plan and performed the data analysis. AP, DD, NJ, ES, MJ, MN, SKB and CSW participated in writing the protocol. All authors read and approved the final manuscript

## Acknowledgements

We would like to thank the adolescents who participated in the study. We would like to extend our sincere gratitude to the Executive Director, Adeodata Kekitiinwa, and the research office; Baylor College of Medicine, Kampala, Uganda, particularly, Dr. Grace Kisitu, Dr. Patricia Nahirya, Ms. Sarah Nakalanzi. We would also like to thank Dr. Jackline Barungi for assisting in patient referral.

